# Government to Government (G2G) Model for HIV Service Delivery – An Approach for Program Ownership and Sustainability in Four Provinces of Zambia

**DOI:** 10.1101/2025.04.08.25325437

**Authors:** Keith Mweebo, Annie Mwila, Kiva F. Tinglof, Mark W. Tenforde, Brian Muyunda, Gregory Parent, Maybin Mumba, Thomas Mupashi, Diane Morof, Jennifer Nel, Leoda Hamomba, Nzali Kancheya, Linos Mwiinga, Eugenia Zulu, Carlistus Kaayunga, Mathews Ngambi, Simulyamana A. Choonga, Lloyd Mulenga, Michele Montandon, Brittney Baack, Steven Y. Hong, Isaac Zulu, Genessa Giorgi, Andrew Auld, Simon Agolory

## Abstract

**Introduction:** Zambia has achieved ambitious programmatic targets of HIV diagnosis, treatment coverage, and viral load suppression among persons living with HIV (PLHIV) with the support of the United States President’s Emergency Plan for AIDS Relief (PEPFAR) and other donors. Beginning in fiscal year (FY) 2019, U.S. CDC Zambia transitioned its PEPFAR Direct Service Delivery (DSD) support in Eastern, Lusaka, Southern, and Western Provinces from non-governmental organizations (NGOs) to a government to government (G2G) provincial health office (PHO) service delivery model for program ownership and sustainability. We reviewed programmatic and financial data during and after the period of this transition.

**Methods:** Programmatic performance using PEPFAR’s Monitoring, Evaluation, and Reporting and expenditure reporting data were used to assess changes over the period of transition from an NGO-led to government-led DSD model in Zambia. Data were reviewed across six FYs from October 2018 – September 2019 (FY19) through October 2023 – September 2024 (FY24) in four provinces across Zambia. Programmatic and expenditure performance indicators were analysed across time to assess PLHIV on antiretroviral therapy (ART), viral load coverage and suppression, and costs associated with care and treatment services.

**Results:** From FY19 to end of FY24, the number of PLHIV on ART across the four provinces increased by 31% (520,628 to 680,781). Viral load coverage and suppression in the transition provinces increased from 73% to 88% and 91% to 98%, respectively. Annual PEPFAR financial investments for care and treatment activities (in 2019 US dollars) increased by 25% ($12.7 million) from $38.3 million in FY19 to $51.0 million in FY21, as the amount invested in DSD increased concurrently with investments to implementing partners. From FY21 to FY24, spending declined by 41% ($21.1 million), while the total number of PLHIV on ART increased. The amount spent per PLHIV on ART decreased from $79 in FY21 to $44 in FY24, a decline of 44%.

**Conclusions:** The G2G model of service delivery and funding is an efficient and sustainable model for HIV epidemic control in Zambia. This government-led model shows a path forward to achieving sustainable HIV program success, reductions in costs, and expanded government ownership of the HIV program.

## Introduction

The global HIV response has reached a point where the UNAIDS goal of ending AIDS as a public health threat by 2030 is a possibility (1). This is demonstrated by the lowest annual number of new HIV infections in 2023 (1.3 million, representing a 39% reduction from 2010) since the epidemic began. Additionally, 30.7 million (79%) of the estimated 39 million people living with HIV (PLHIV) globally are accessing life-saving antiretroviral therapy (ART) (2). Sub-Saharan Africa has made the most progress towards achieving the UNAIDS 2030 goal, with marked reductions in the number of new HIV infections and AIDS-related deaths (a 56% reduction in number of new infections and deaths since 2010). The global HIV response is primed to transition from an emergency to a sustained response that maintains the treatment cohort on life-saving drugs and viral suppression (HIV viral load < 1000 copies/ml). Changing current approaches to focusing on closing remaining gaps and ensuring programs are led and delivered by national governments and other domestic entities including communities are key elements of the transition from an emergency response to a sustained approach (2).

Zambia has contributed to Sub-Saharan Africa’s successful progress towards ending AIDS as a public health threat by 2030. UNAIDS estimates that, of the 1.3 million Zambians living with HIV in 2023, 1.2 million were on life saving treatment and had suppressed HIV viral loads. Based on these UNAIDS estimates, 96% of Zambians living with HIV know their HIV status and 97% were on ART, with a population-level viral load suppression (VLS) of 92% among PLHIV as of December 2023 (3). These 2023 UNAIDS estimates suggest that progress has been made since the most recent Zambia Population-Based HIV Impact Assessment (ZAMPHIA) survey in 2021, which found that 89% of Zambians living with HIV knew their status, 98% of those who knew their status were on treatment, and 96% of the treatment cohort was virally suppressed, corresponding with a population-level VLS of 86% for PLHIV (4).

Historically, the President’s Emergency Plan for AIDS Relief (PEPFAR)-funded HIV programs that have supported HIV services in Zambia and other countries over the past two decades have been implemented through direct funding to international non-governmental organizations (NGOs) and academic institutions who set up programs and hired technical experts from the United States. and other countries. This was the initial approach as part of the emergency response to the HIV epidemic partly due to limited or lack of expertise to support HIV programing in many resource-limited settings. There are significant overhead investments associated with this model, including facilities and administrative costs. A review of international NGO budgets funded by CDC Zambia show these costs can range from 45% to 50% of the total budget of a contract or grant. On the other hand, implementation by local partners and governments is associated with much lower overhead costs. The need for a sustainable model informed the decision to transition from the international NGO-led HIV response to a government-to-government (G2G) model in the four CDC supported provinces in Zambia.

As part of the transition from an emergency HIV response largely led and implemented by international NGOs and academic institutions to a response led by national government and local entities, the U.S. Centers for Disease Control and Prevention’s (CDC’s) PEPFAR-funded program in Zambia started transitioning to a G2G implementation model in fiscal year (FY) 2019 (October 2018–September 2019).

The G2G model is based on support to four out of ten provincial health offices (PHOs) and is one part of a comprehensive HIV program. PHOs work under the authority of the National Ministry of Health (MOH) to implement all health programs in provinces including HIV service delivery. The G2G model utilizes a cooperative agreement (CoAg) funding mechanism for PHOs with substantial engagement of CDC staff in programmatic, technical, human resource, and financial management oversight. The critical role played by CDC’s technical and financial experts working with the PHO ensures accountability and maximizes the impact of every dollar invested.

We aim to describe the programmatic outcomes and financial expenditure during and after the transition of HIV treatment and service delivery from an NGO to a G2G model in Zambia.

## Methods

### G2G Model Implementation

Beginning in FY 2019, using a CoAg grant funding mechanism, CDC began empowering government-led direct HIV service delivery programs by directly funding four PHOs. The G2G model of service delivery is based on provision of funds to PHOs who are mandated under MOH to provide all health services including for HIV and tuberculosis (TB). The PHO is a government subnational health unit that supports health service delivery in districts. Under the government structure, PHOs receive monthly grants from the central government to provide technical assistance (TA) and oversight to district health offices (DHO), that in turn implement health service delivery through health facilities. PEPFAR funding provided to PHOs under the G2G model to implement HIV services is channeled to DHOs and health facilities using the existing government structures.

CDC technical teams engaged in substantial capacity building interventions including trainings and mentorship. The TA provided by CDC technical teams focused on implementation of evidence-based interventions. For sustainability of the TA, the PHOs recruited medical officers and HIV nurse practitioners who were trained as HIV mentors. CDC’s technical experts supported capacity building among all mentors who provided mentorship to frontline health care workers. The mentorship program has since expanded to all provinces in the country. The national mentorship program includes mentorship by national and subnational technical experts through digital platforms under the extension of community health outcomes (ECHO) platform (5). As the program has continued to mature, engagement by CDC technical teams for direct TA to frontline health care workers and subnational mentors has reduced with the TA primarily provided through the Zambian government-led clinical mentorship program (6).

Technical assistance from CDC also included building strong monitoring and evaluation (M&E) systems. Enhanced M&E systems incorporated weekly review of priority indicators in situation rooms, monthly meetings and granular site management focused on site level performance reviews to identify gaps for remediation. The reviews resulted in the generation of action plans that were tracked on a weekly basis. Quarterly performance review meetings included leadership from the MOH headquarters and CDC Zambia office. This forum enhanced leadership buy-in and acted to resolve actions that required senior leadership both from MOH and CDC including policy decisions that helped address key programmatic gaps in a transparent manner with clear roles and responsibilities. Through the funding provided, PHOs hired additional clinical, laboratory, pharmacy and M&E staff and supported community-based volunteers to fill critical human resources gaps.

Additionally, PHOs procured vehicles to facilitate transportation of district supervisor/mentorship teams to health facilities and support timely distribution of commodities and samples for various laboratory tests, procured equipment needed for service delivery, and provided financial resources to districts to support the direct service delivery (DSD) efforts at health facilities and communities.

To enhance capacity to manage financial resources and improve technical competencies of PHO and DHO staff, CDC’s finance and grants management office provided trainings and mentorship on financial management and compliance to United States government (USG) funding regulations as well as Zambian government financial requirements. Monitoring of compliance to regulations was done including comprehensive assessments like business process reviews (BPR). A BPR is a process that evaluates an organization’s operations and assesses its current processes, procedures, and systems in place for managing the CoAg to ensure that they are efficient, effective, and in compliance with terms and requirements by USG regulations, host country regulations and legislation, and industry good practice.

In the first year, all districts in Eastern Province, three districts out of five in Lusaka Province, five districts out of 13 in Southern Province, and three districts out of 13 in Western Province transitioned from an NGO-based model of HIV service delivery to a G2G model. All the remaining districts transitioned to the G2G model in FY20 (Figure 1).

**Figure 1.**
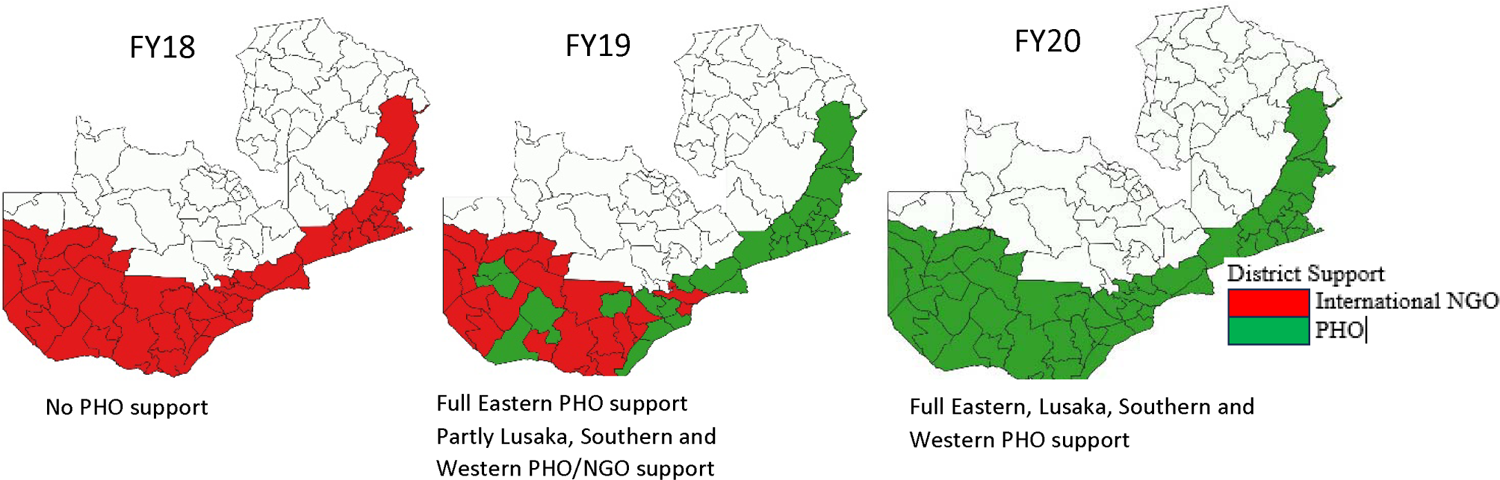
Phased approach from an NGO-led to G2G model of HIV service delivery program implementation over time – four provinces in Zambia, FY18 to FY22. Abbreviations: FY = fiscal year, G2G = government-to-government, NGO = non-governmental organization, PHO = Provincial Health Office

Since FY20 (October 2019–September 2020), all four CDC-supported provinces have transitioned from the NGO model to a G2G model for delivering services through PHOs. PHOs were responsible for defining the programmatic needs, including human resources and technical and administrative oversight, to meet adult and pediatric HIV prevention, care and treatment targets. When needed, both international and local TA partners provided additional targeted support to assist the PHO to achieve their programmatic and service quality priorities. For example, if a province needed additional technical support to achieve their VLS rates among children, PHOs work with the TA partner to identify evidence-based interventions that can be implemented by the province. Once a PHO demonstrated readiness through consistent achievement of targets, strong technical partnership with CDC, and effective business management processes, the TA partner’s scope was narrowed/reduced and eventually phased out. This phased approached was assessed across the 6 years of implementation to better characterize gains observed through transitioning to the G2G-led model. Southern Province was the first province to phase out TA in FY24.

### Study design

We analyzed HIV program data from the four provinces in Zambia (Eastern, Lusaka, Southern, and Western) where the G2G model was implemented. Program performance was assessed over six FYs from October 2018–September 2019 (FY19) through October 2023–September 2024 (FY24). Additionally, PEPFAR financial investments trends under the care and treatment budget in the 6-year period were analyzed and compared to results recorded for the treatment cohort. We excluded program data from one academic teaching hospital (University Teaching Hospital) in Lusaka Province supported under a separate CoAg with CDC during FY19–FY24 as well as community-based treatment and prevention projects in Lusaka Province supported by the United States Agency for International Development (USAID) during the G2G period (Zambia DISCOVER-Health Project in FY19–FY20 and Controlling HIV Epidemic for Key and Underserved Populations in FY23–FY24). These were excluded because the scope of work was not exclusive to the areas under the G2G model in the four provinces and financial data for care and treatment at provincial level were not available.

### Data sources

PEPFAR program and financial data were reviewed over the 6 years since the program transitioned to G2G support in the four CDC-supported provinces in Zambia. We focused on program performance measures related to service delivery to assess the G2G transition. We used PEPFAR Monitoring, Evaluation, and Reporting (MER) data to assess HIV clinical service outcomes and expenditure reporting (ER) data to assess financial contributions to program implementation.

### Indicators/measures

To assess HIV clinical service delivery during G2G model implementation across the four CDC-supported provinces, we used MER indicator data to evaluate key performance indicators. We focused on routine HIV clinical services measuring HIV treatment data and viral load monitoring. MER data reporting processes have been described previously (7). Clinical indicators of interest for this analysis included: 1) the absolute number of PLHIV currently on ART, defined as the number of PLHIV on ART at the end of the reporting period; this excluded individuals who did not receive antiretrovirals within 4 weeks of their last scheduled medication pick-up; 2) the percentage of PLHIV on ART who received routine viral load testing at 6 months post initiation and every 12 months thereafter based on national guidelines, proxy (viral load coverage [VLC]) defined as the percentage of eligible individuals on ART with a valid viral load result documented in the medical or laboratory records / laboratory information systems within the past 12 months (8); and 3) of PLHIV on ART who received viral load testing, the percentage of these individuals who achieved VLS. VLS is defined as the percentage of individuals on ART with a suppressed viral load result (<1,000 copies/ml) documented in the medical or laboratory records/laboratory information systems within the past 12 months. We described these aggregate HIV performance indicators at the end of each FY, overall and stratified by province, to assess trends in care and treatment performance indicators over time.

ER data were used to evaluate financial resource requirements in shifting from an NGO to a G2G approach. Under ER, PEPFAR collects all expenditures annually from all implementing partners that receive PEPFAR funds. Implementing partners report on a standardized classification system that collects expenditures by program area, beneficiary group, and cost categories. This analysis focused on the care and treatment (C&T) and program management (PM) program areas. C&T includes any spending related to the delivery of services and TA related to the care and treatment of PLHIV on ART, including all related laboratory services. PM includes any spending related to coordination and support to administration of the program, usually under management and operations of the grant. Using ER allows us to evaluate the shift in input requirements in moving to G2G on the total financial requirements, in addition to shifts in investment footprint (e.g. spending on service delivery implementation relative to overhead investments). Further, ER input indicator data are paired with MER output indicator data to develop process indicators that explore the impact on efficiency. To remove the effect of inflation on resource requirements, constant 2019 US dollars were used in this analysis. Finally, the impact of programmatic shifts on overhead costs are evaluated by reviewing overhead cost ratio (spend on program management divided by total spend) and overhead spend per output (total spend divided by PLHIV on treatment). These approaches are commonly used in the private sector as indicators for efficiency of operations.

### Data analysis

All analyses were descriptive, with results stratified by year. Microsoft Excel (Redmond, Washington, USA) was used to perform analyses.

### Ethics

All data reported in this analysis were de identified, aggregate, routine HIV program data collected as part of PEPFAR support in Zambia. This activity was reviewed by CDC, deemed research not involving human subjects, and was conducted consistent with applicable federal law and CDC policy.^§^

## Results

### MER Clinical Indicator Performance

Program data reported from FY19 (October 2018–September 2019) to the end of FY24 (October 2023–September 2024) indicate the cohort of PLHIV on ART grew overall, with a growth observed in the treatment cohort across all four G2G-supported provinces and year-to-year growth in consecutive years except FY22 (Figure 2A). From FY19 to end of FY24, the number of PLHIV on ART across provinces increased by 37%, from 495,552 to 680,781 PLHIV. This ranged from a 25-48% growth in the treatment cohort size from FY19 to FY24 across the four provinces.

**Figure 2.**
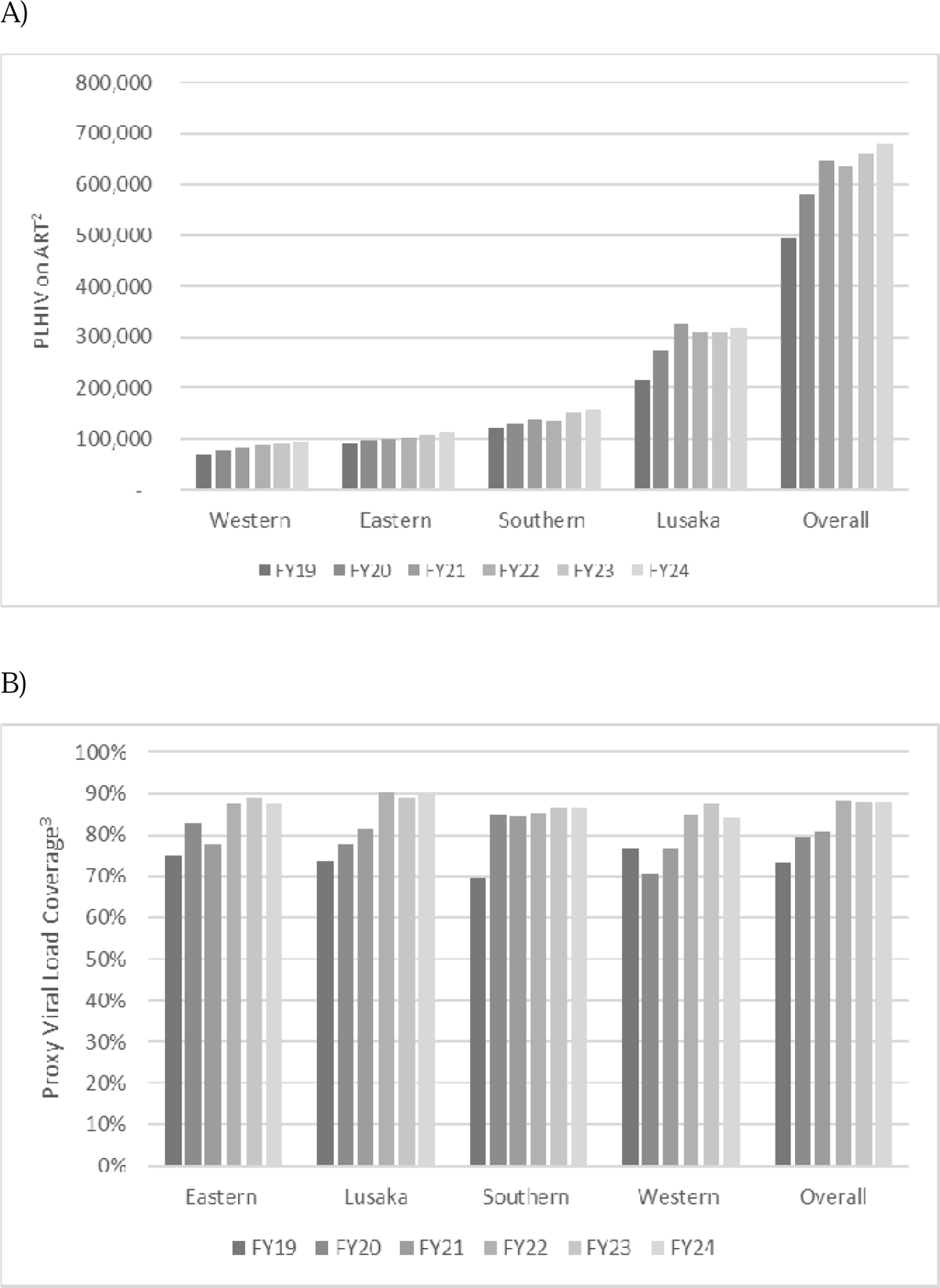
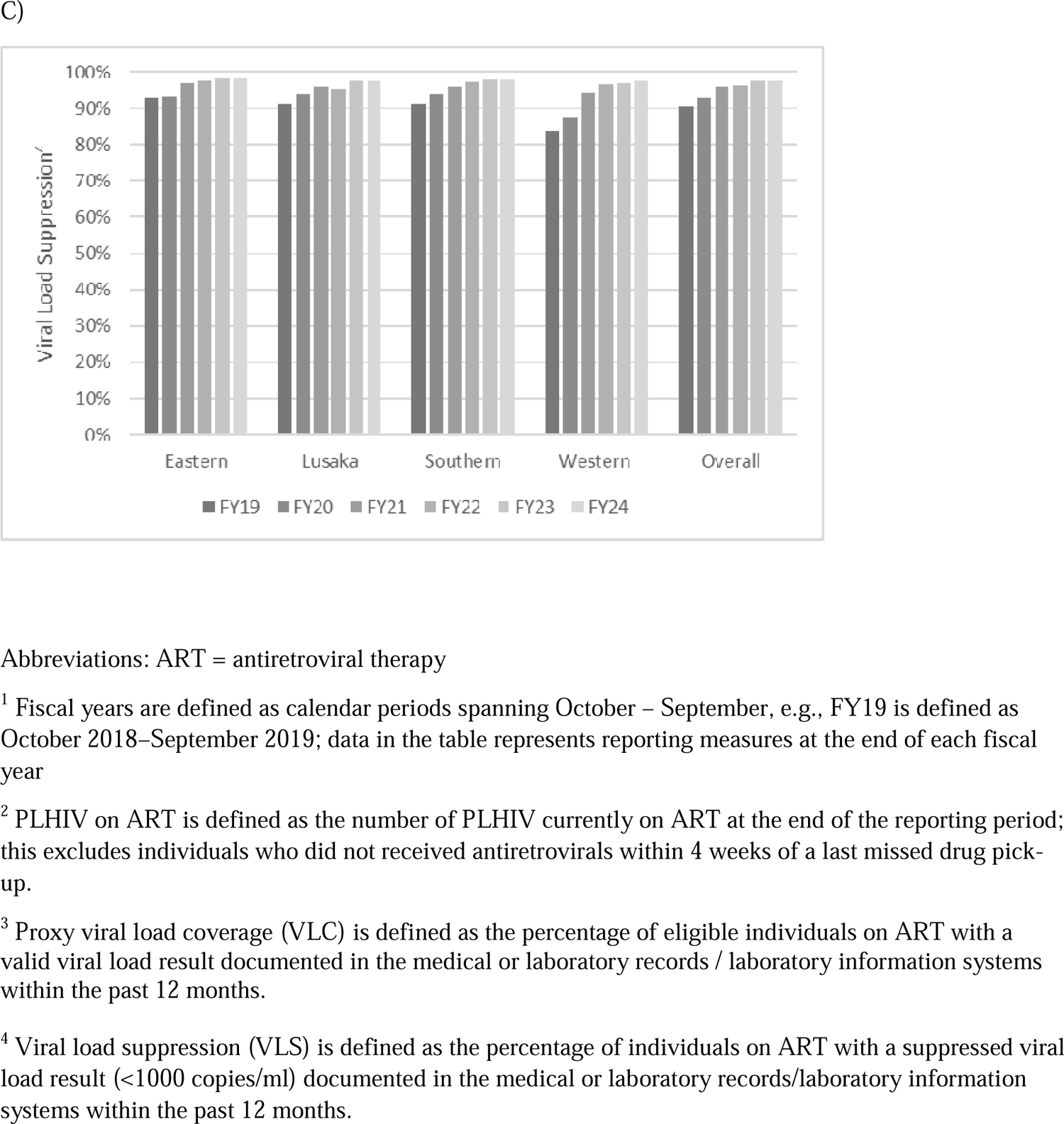
Programmatic HIV service delivery outcomes for A) number of PLHIV on treatment, B) VLC and C) VLS in four PEPFAR-supported provinces in Zambia by fiscal year (FY)^1^, October 2018 to September 2024

VLC and VLS outcomes showed improvements over the 6-year period following implementation of the G2G model (Figure 2B, Figure 2C). VLC increased from 73% in FY19 to 88% in FY24, with VLC reaching 88% during the final three FYs (Figure 2B). Among PLHIV on ART who received viral load testing, the percentage of these individuals who achieved VLS also increased, from 91% to 98% during the last two FYs (Figure 2C). Improvements in viral load measures were observed across all provinces.

Financial investments for care and treatment activities showed an increase from FY19 to FY21 from $38.3 million to $51.0 million (Figure 3A). After the first 2 years of G2G implementation, costs declined each year with a cost total of about $30 million in FY24.

**Figure 3.**
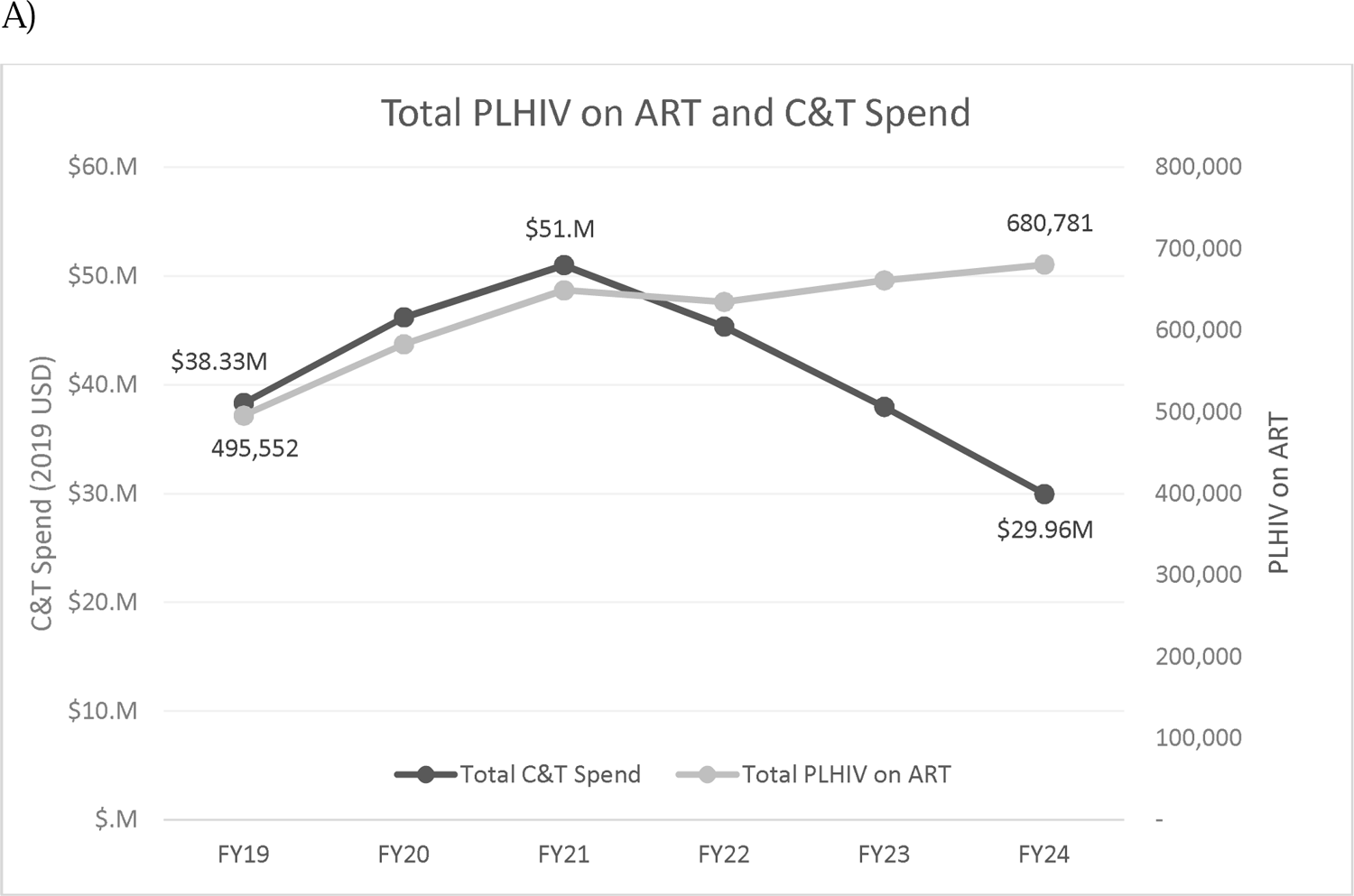
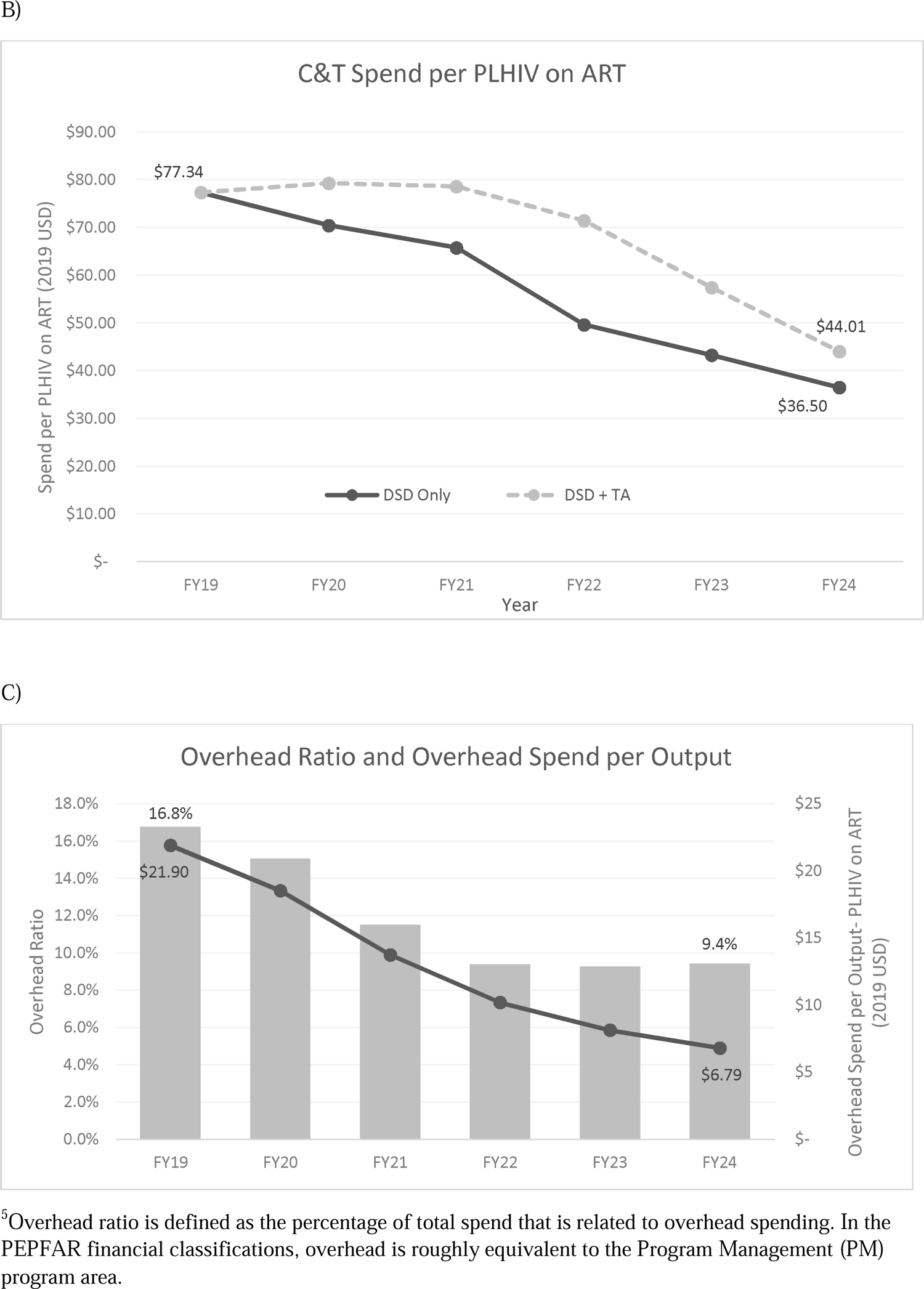
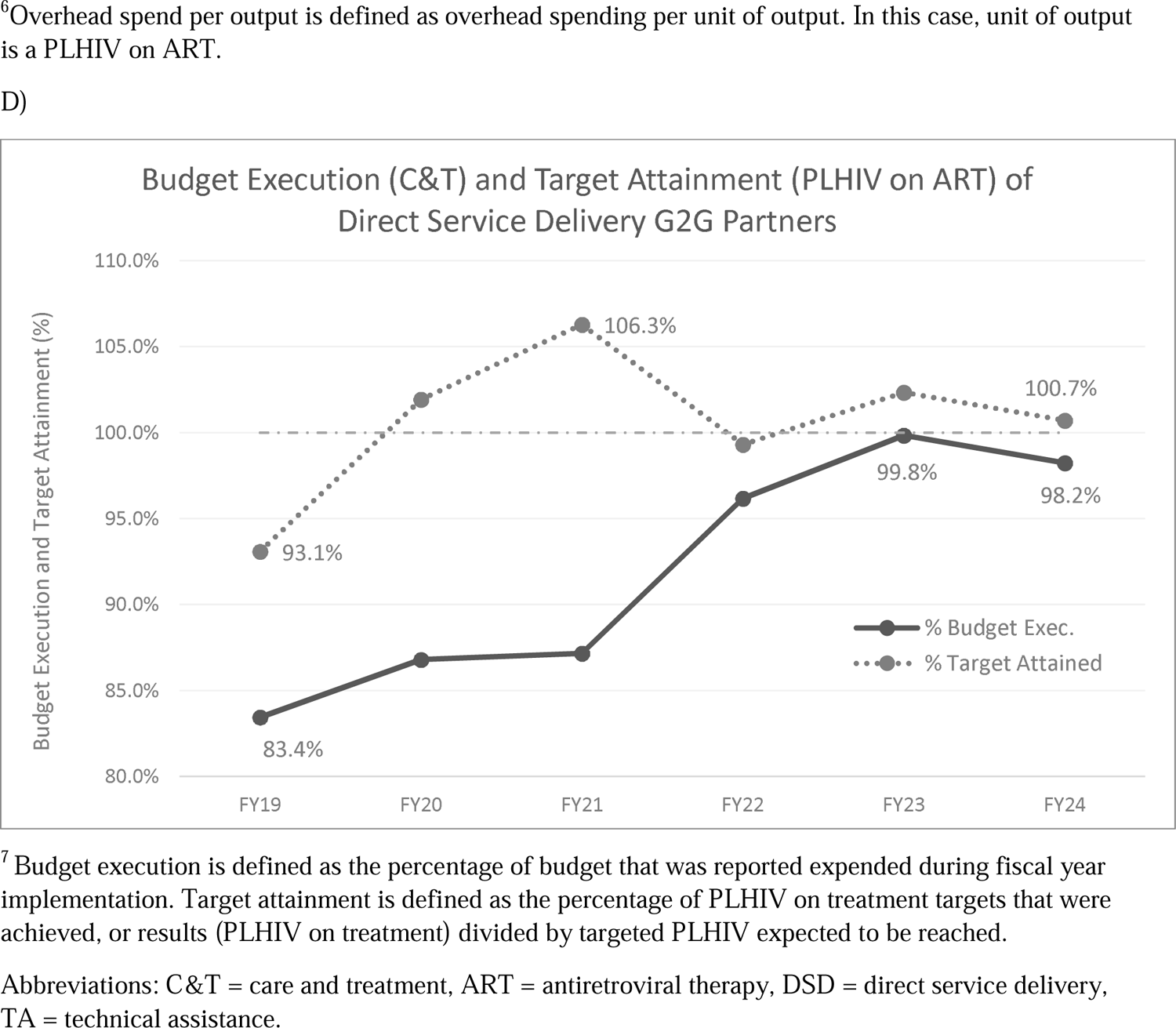
Programmatic HIV service delivery outcomes and expenditure for A) Total people living with HIV (PLHIV) on antiretroviral therapy (ART) results and amount spent for care and treatment activities in US dollars (USD), B) Amount spent on care and treatment (C&T) activities including direct service delivery (DSD) and technical assistance (TA) per PLHIV on ART, C) Overhead Ratio and Overhead Spend per Output (PLHIV on ART), D) Care and treatment budget execution and target achievement for PLHIV on ART,

As the DSD activities transitioned from NGO-led to a G2G approach in the provinces, total spending on C&T activities initially increased by $12.7 million or 25% (2019 USD) from FY19 to FY21 as the amount invested in DSD increased concurrently with investments to implementing partners providing TA to the provincial G2Gs (Figure 3A and 3B). The total PLHIV on ART also increased roughly similar to the spending, with a 24% increase observed. This resulted in a flat total spend per PLHIV from FY19 through FY21. However, from FY21 to FY24, total C&T spending declined by 41% ($21.0 million), while total PLHIV on ART increased by 4.9% (31.6K) (Figure 3A). This led to a 44% decline in C&T spend per PLHIV on ART from $79 in 2021 to $44 in 2024 (Figure 3B). In looking at total change in C&T spend per PLHIV on ART from 2019 to 2024, DSD-only partner spend declined by $41 (53%), while DSD and TA partner spend declined by $33 (43%).

Figure 3C illustrates that shifting DSD to provincial G2G partners has reduced overhead spending. The overhead spend ratio of service delivery in FY19 was 16.8% and declined to 9.4% in FY24. Additionally, the overhead spend per PLHIV on ART declined from $21.90 in FY19 to $6.79 in FY24, a decline of over 68%. Budget execution of C&T programs increased from 83.4% to 98.2%, while attainment of program targets for PLHIV on ART during each FY improved from 93.1% to >100% (Figure 3D).

## Discussion

This review of HIV program and expenditure data shows that Zambia’s transition from an NGO to a G2G model of HIV treatment service delivery has been impactful, programmatically sustainable, and financially efficient. Results from this analysis suggest that indicators of PLHIV receiving ART continue to perform well after transitioning activities to a government-led model of delivering HIV services across four provinces in Zambia. During the transition from an NGO-to G2G model, financial investments per PLHIV on treatment decreased and allocation of dollars and the amount spent during a FY were more closely aligned (i.e., budget execution improved), all while program gains were observed in the number of PLHIV on treatment across G2G provinces. The impact, efficiency, and sustainability of Zambia’s HIV treatment program following transition from an NGO-led to a host government-led model of service delivery can be relevant for other countries in the region and globally. Findings from this analysis can help design roadmaps to sustainability centered around transitioning HIV programs towards government leadership and management of the response (9, 10).

During the 6-year period while the G2G model was initiated and scaled up in the four provinces, stable increases were observed in the cohort of individuals receiving ART. Further, viral load indicators also saw improvement over time across provinces, demonstrating that these provinces effectively maintained patients in care with routine laboratory monitoring and that patients had a sufficient level of ART adherence with most obtaining VLS. Although this analysis only focused on the four out of ten PEPFAR-supported provinces in Zambia that transitioned from an NGO service delivery model to a government-led model, other representative population-based surveys also support quality of HIV services with the G2G model. For example, the 2021

Zambia Population-based HIV Impact Assessment (ZAMPHIA) survey found VLS ranging from 88-93% among PLHIV aged ≥15 years in G2G-supported provinces compared with 86% across all ten provinces (4). Additionally, although based on only one year, quality of services – as indicated by key performance indicators assessed – was maintained despite graduation from TA partner support in one of the four provinces (Southern).

The initial increase in financial investment under care and treatment in the first 2 years were driven by ART surge campaigns to accelerate closing of the treatment gaps. These efforts sought to ensure the program was on track to achieve the UNAIDS 95-95-95 goals by 2025 and the continued investments in TA partners. TA activities focused on optimizing case identification, linkage to treatment, and improve treatment coverage. The noted reduction in overhead spend ratio of service delivery from 16.8% in FY19 to 9.4% in FY24 suggests that the G2G approach in Zambia has been a cheaper service delivery model. This is important for sustainability as local governments and entities will have less resources available to support the HIV response beyond epidemic control. Using PLHIV on ART as the program output, the overhead spending per PLHIV on ART declined by over 68%. The financial metrics reviewed during the transition period indicate an improvement in the efficiency of service delivery implementation of PEPFAR resources through the G2G model.

A recent analysis showed that over the four-year period, high-risk BPR findings fell from 88 to 14 (an 84% reduction) and monetary audit findings fell from 20 to 4 (an 80% reduction) (11). The improvement in adherence to regulations noted in the publication aligns with the improvement in budget execution recorded in this analysis beginning in FY21. During this period, CDC Zambia was able to increase direct funding to the PHOs by 300% from $13 million to $52 million. With the funding increase, the PHOs expanded healthcare staff positions by 259% from 2,621 to 9,397 (11). The BPR and direct capacity building of PHOs in financial management have been part of the transition from an NGO-led to G2G model, which increased the government’s ability to receive PEPFAR funds and resulted in significant HIV program achievements. Evidence suggests that transitioning to government-led service delivery in Zambia has been achievable when BPRs, internal control assessments, and proactively mitigating risks are integrated in the transition process and program strategy (11).

In FY24, Southern Province graduated from TA support and continues to maintain programmatic gains across HIV clinical indicators (4). Additionally, the province demonstrated good financial management based on findings of BPR conducted in 2024. Review of FY24 program data indicate that more provinces achieved the programmatic 95-95-95 UNAIDS targets. Findings of BPR reviews for financial year ending September 30, 2024, will further inform graduation from TA support from the remaining three provinces. Analysis in future years will be important to evaluate whether program success is sustained in provinces that graduate from TA support.

This study was subject to several limitations. First, HIV programmatic and ER data are collected in aggregate quarterly and annually, respectively, and can vary in data quality and completeness. Second, the contributions of individual activities to improving HIV service delivery performance measures and generating efficiency were not systematically implemented across provinces and therefore could not be quantified. Third, there was no comparison done with provinces not implementing the G2G model. Also, we did not include CDC staff time for engagement on mentorship and training on financial management and compliance, and programmatic support and TA.

## Conclusions

As HIV programs begin to reach the UNAIDS goal of 95-95-95 and shift from an emergency to a sustained response, efforts to find sustainable and efficient program models can be drivers of successful public health efforts. The G2G model provides an example for how CoAg structures can support government-led HIV programs that sustain gains and promote sustainable implementation of activities. Our findings show that the G2G model was an efficient and sustainable model for HIV service delivery in Zambia.

## Competing interests

The authors have no funding or conflicts of interest to disclose.

## Ethical footnote

^§^See e.g., 45 C.F.R. part 46; 21 C.F.R. part 56; 42 U.S.C. §241(d), 5 U.S.C. §552a, 44 U.S.C. §3501 et seq.

## Authors’ contributions

KM served as the lead author and supported overall manuscript planning, development and participated in defining data elements, review, writing of results and interpretation. AM supported and led the implementation of the G2G model and participated in the initial planning and conceptualization of the manuscript. KT supported manuscript planning, writing, coordination of all authors, led review of journal requirements and procedures for adhering to publication. MWT participated in manuscript development especially methodology, data analysis, writing of results and discussion. Also participated in development review meetings and auditing. DM supported overall manuscript conceptualization, planning, development, and writing. LM supported manuscript review and provided context to implementation in the early years of the G2G model. BM contributed to methods, data analysis and added context to early years of G2G implementation. MM conducted MER data analysis and validation of data and implementation road map. GP conducted ER data analysis and development of methods and writing of results. Also participated in planning of the manuscript and conducted audit and reviews. JN conducted MER data validation critical review of the manuscripts. TM supported capacity building of PHO during implementation with adherence to financial regulations and led the analysis of business review processes. He also contributed to writing of the manuscript. EZ supported capacity building of PHO during implementation with adherence to financial regulations and led the analysis of business review processes. LH supported review of HRH data and was point of contact during the scale up phase of the model and provided key guidance on HRH needs at service delivery points. NK supported manuscript planning and development and ensured adherence to science processes and supported implementation of the G2G process during her previous role on programmatic work. CK, MN &AC were provincial leads from the government during implementation and provided guidance to program and financial management under the G2G. LM provided oversight guidance to the PHOs and supported programing and reviewed the manuscript for adherence to Ministry of health procedures. IZ provided TA to CDC country office team during early days of implementation. MM, BB, and SH provided TA to the country team on implementation and manuscript planning and data analysis. AA supported implementation of G2G and part of the team that planned for systematic analysis of the G2G results. SA and GG supported with the initial conceptualization of the G2G model and supported implementation.

## Data Availability

All data produced in the present work are contained in the manuscript.

## Acknowledgements

Rachel Johnson, CDC Zambia country Office of Director for her support during data analysis and writing of the manuscript. Zambia Ministry of Health.

## Funding

This publication has been supported by the U.S. President’s Emergency Plan for AIDS Relief (PEPFAR) through the U.S. Centers for Disease Control and Prevention (CDC).

## Disclaimer

The findings and conclusions in this manuscript are those of the authors and do not necessarily represent the official position of the funding agencies.

## Notes

### Competing Interest Statement

The authors have declared no competing interest.

### Funding Statement

This publication has been supported by the U.S. President's Emergency Plan for AIDS Relief through the U.S. Centers for Disease Control and Prevention.

### Author Declarations

This activity was reviewed by the U.S. Centers for Disease Control and Prevention (CDC), deemed research not involving human subjects, and was conducted consistent with applicable federal law and CDC policy.

## References

1. Joint United Nations Programme on HIV/AIDS. The urgency of now: AIDS at a crossroads. 2024;CC BY-NC-SA 3.0 IGO.

2. U.S. State Department. PEPFAR Program Strategy Discussion Guidance. 2024.

3. UNAIDS country fact sheets: Zambia [Internet]. 2024. Available from: https://www.unaids.org/en/regionscountries/countries/zambia.

4. Mulenga LB, Hines JZ, Stafford KA, Dzekedzeke K, Sivile S, Lindsay B, et al. Comparison of HIV prevalence, incidence, and viral load suppression in Zambia population-based HIV impact assessments from 2016 and 2021. AIDS. 2024;38(6):895–905.

5. The University of New Mexico. Project ECHO 2023 Annual Report. 2023.

6. Boyd MA, Fwoloshi S, Minchella PA, Simpungwe J, Siansalama T, Barradas DT, et al. A national HIV clinical mentorship program: Enabling Zambia to accelerate control of the HIV epidemic. PLOS Glob Public Health. 2022;2(2):e0000074.

7. Data for Accountability TaI. Monitoring, Evaluation, and Reporting Indicator Reference Guide 2024 [Available from: https://help.datim.org/hc/en-us/articles/360000084446-MER-Indicator-Reference-Guides.

8. Zambia Ministry of Health. Zambia Consolidated Guidelines for the Treatment and Prevention of HIV. 2022.

9. U.S. State Department. PEPFAR Five-year Strategy 2022 [Available from: https://www.state.gov/pepfar-five-year-strategy-2022.

10. UNAIDS – planning for sustainability of the HIV response up to and beyond 2030. [Internet]. 2024. Available from: https://www.unaids.org/en/resources/presscentre/featurestories/2024/april/20240430_sustainability.

11. Mupashi T, Zulu E, Chona Y, Hamomba L, Mateyo T, Muyunda B, et al. Sustainable Business Process Model for Achieving Excellence in HIV Service Delivery through Government to Government (G2G) Funding and Direct Capacity Building of Provincial Health Offices in Zambia. INTEREST Scientific Conference2023.

